# Verification of Self-Reported HPV Vaccination in the Initial Cohorts of the Botswana’s National Immunization Programme, 2023–2024

**DOI:** 10.64898/2026.06.23.26355877

**Authors:** Doreen Ramogola-Masire, Anikie Mathoma, Malebogo Masono, Maitumelo Masole, Sibongile Phiri, Jared Omolo, Rayleen M. Lewis, Chelsea Morroni, Rebecca Luckett, Lauri E. Markowitz, Julia W. Gargano

**Affiliations:** Department of Obstetrics & Gynecology, University of Botswana, Gaborone, Botswana; Division of Research & Enterprise, University of Botswana, Gaborone, Botswana; CDC Botswana, Gaborone, Botswana; CDC, National Center for Immunization and Respiratory Diseases, Division of Viral Diseases, Atlanta, GA, USA; Centre for Reproductive Health, University of Edinburgh, Edinburgh, United Kingdom; Botswana Harvard Health Partnership, Gaborone, Botswana; Beth Israel Deaconess Medical Center, Department of Obstetrics and Gynecology, Boston, USA

**Author notes:** **Disclaimer:** The findings and conclusions in this report are those of the author(s) and do not necessarily represent the views of the US Centers for Disease Control and Prevention.

## Abstract

Botswana launched a national female-only school-based 2-dose HPV vaccination programme among 9–13-year-olds in 2015. During a 2023–2024 vaccine impact study among women aged 18–22 years recruited from the University of Botswana and Gaborone-area HIV clinics, we compared self-reported HPV vaccination to school and health facility records. Of 450 women, 446 self-reported HPV vaccination (301: 1-dose, 135: 2-dose, 10: 3-dose). Vaccination status was verified for 429/450 (95%); one was not vaccinated, 52 (12%) had 1 dose, 344 (80%) 2 doses, and 32 (7%) 3 doses. Agreement on ≥1 dose was >99%; dose-specific agreement was 37% (159/429); 256 (60%) underreported and 14 (3%) overreported number of doses. While time- and resource-intensive, nationwide vaccination verification without a vaccination registry was feasible. Results suggest high vaccination coverage, including high ≥2-dose coverage. Self-report of ≥1 dose was highly accurate, but number of doses was often underreported.

## INTRODUCTION

Botswana is an upper middle-income country in Southern Africa with a population of 2.45 million people. In Botswana, human immunodeficiency virus (HIV) prevalence among women aged 15–49 years was 17.9% in 2022 (1), and cervical cancer is the most common cancer (2). Most cervical cancers are caused by vaccine-preventable human papillomavirus (HPV) types 16 and 18; women living with HIV (WLHIV) bear a disproportionate burden of diseases caused by HPV, including cervical cancer (3, 4, 5). In 2013, Botswana conducted a demonstration project of 3-dose HPV vaccination of girls in one community, achieving high coverage (83% ≥1-dose) (6). Botswana then launched a national school-based 2-dose HPV vaccination programme in 2015 in a multi-age cohort of 9–13-year-olds; from 2016 onward, the national routine immunization programme in Botswana focused on HPV vaccination of girls aged 9 years, equivalent to grade 5 in school.

The University of Botswana (UB) and the US Centers for Disease Control and Prevention (CDC) evaluated the impact of Botswana’s HPV vaccination programme through two cross-sectional studies of HPV prevalence among persons aged 18–22 years. In the first study (2019– 2020), participants were not age-eligible for the national vaccination program. In the second study (2023–2024), most female participants were age-eligible when the program started. We aimed to verify vaccine-eligible female participants’ HPV vaccination history, including number of doses received and vaccination dates, through information recorded at the time of vaccination.

## MATERIALS & METHODS

All 2023–24 study participants were asked whether they had received HPV vaccination and related details (e.g., residential location and primary school when vaccinated) during a structured interview with a research assistant. Female and male participants were asked because although only females were included in Botswana’s vaccination programme, some males could have received vaccination through the private sector. We sought to verify the HPV vaccination history for all female 2023–2024 study participants only.

We piloted vaccination verification procedures for 27 participants in Gaborone. Through this pilot, we determined that vaccination records could be located for most participants. Research assistants visited primary schools that women reported attending and/or health facilities that served their schools or communities to find and review vaccination records. Following this pilot, using residential location and primary school attended, research assistants carried out additional visits in and around Gaborone and travelled throughout Botswana for record verification. Research assistants traveled to each location by car or plane and stayed 1–3 weeks in a city, town, or large village to search for vaccination records in the vicinity.

During the verification visit, date and location of vaccination (school, health facility, other) and source of data (vaccination card, school record, health facility record) were recorded for up to three vaccine doses. Participants who had ≥1 HPV vaccine dose documented with the date administered were considered verified as vaccinated. Participants were considered verified as not vaccinated if school records corroborated the student’s attendance on the date of vaccination and neither school nor health facility records included a vaccination record, or if school or health facility records indicated the student was not vaccinated, e.g. due to parental refusal.

Descriptive analyses were conducted overall and by group, i.e. women without HIV, WLHIV overall, WLHIV whose HIV infection was perinatally acquired (WLHIV-PA), and WLHIV whose HIV infection was not perinatally acquired (WLHIV-nPA). The numbers and proportions with verified data, the cross-tabulation of self-reported and verified doses among those with verified status, the percent agreement of ≥1 dose, and the percent agreement by number of doses were calculated.

This study was reviewed and approved in accordance with the CDC human research protection procedures and approved as human subjects research (protocol 7451.0, accession number NCIRD-HPV-1/26/23-9eb40). It was also approved by institutional review boards at UB and the Botswana Ministry of Health.

## RESULTS

The 2023–2024 study included 500 participants from UB (250 females, 250 males), and 200 WLHIV from Gaborone-area ART clinics. All 250 UB female participants reported receiving ≥1 HPV vaccine dose. Of 200 WLHIV, 195 reported receiving ≥1 HPV vaccine dose. Of 250 UB male participants, 244 reported being unvaccinated, and 6 did not know.

During the vaccination verification process, research assistants determined that vaccination records were easiest to obtain from health facilities. School visitor logs documented the dates of school health nurse vaccine administration visits, but not individual student vaccinations. Doses were recorded on individual HPV vaccination cards; study participants generally did not have these cards, but they were sometimes found in primary school records. Individual vaccination documentation was also found written directly in school health folders. Another source of verification was ledgers kept by health facility nurses, who were responsible for visiting schools and administering vaccines. These contained all vaccine doses given to individual students during school visits.

Records sufficient to verify vaccination were obtained for 429/450 (95%) participants, including 247/250 (99%) women without HIV (UB group) and 182/200 (91%) WLHIV (Table 1). No verification visit was made for 20 women because they attended primary school in another country (n=17) or in a remote area of Botswana (n=3). For one participant, a visit was made, but records were insufficient to verify vaccination status. A lower proportion of WLHIV had verified data than women without HIV (91% vs. 99%). WLHIV-nPA had the lowest proportion verified (88%); among WLHIV-PA, 96% were verified.

**Table 1.** Self-reported and verified vaccination status of women aged 18–22 years participating in an HPV vaccine impact study in Botswana, 2023–2024.

|  | Self-reported doses | Not verified* | Verified Doses |  |  |  | Verified data n/N (%) | ≥1-dose agreement n/N (%) | Dose-specific agreement n/N (%) |
| --- | --- | --- | --- | --- | --- | --- | --- | --- | --- |
|  |  |  | 0 | 1 | 2 | 3 |  |  |  |
| **All women** |  |  |  |  |  |  |  |  |  |
| Not vaccinated by self-report | 4 | 2 | 1 | 0 | 1 | 0 | 429/450 | 428/429 | 159/429 |
| 1 self-reported dose | 301 | 16 | 0 | 43 | 227 | 15 | 95.3% | 99.8% | 37.1% |
| 2 self-reported doses | 135 | 3 | 0 | 8 | 111 | 13 |  |  |  |
| 3 self-reported doses | 10 | 0 | 0 | 1 | 5 | 4 |  |  |  |
| Verified status (total) | 450 | 21 | 1 | 52 | 344 | 32 |  |  |  |
| **Women without HIV** |  |  |  |  |  |  |  |  |  |
| Not vaccinated by self-report | 0 | 0 | 0 | 0 | 0 | 0 | 247/250 | 247/247 | 102/247 |
| 1 self-reported dose | 150 | 2 | 0 | 21 | 125 | 2 | 98.8% | 100% | 41.3% |
| 2 self-reported doses | 93 | 1 | 0 | 5 | 79 | 8 |  |  |  |
| 3 self-reported doses | 7 | 0 | 0 | 0 | 5 | 2 |  |  |  |
| Verified status (total) | 250 | 3 | 0 | 26 | 209 | 12 |  |  |  |
| **Women living with HIV** |  |  |  |  |  |  |  |  |  |
| Not vaccinated by self-report | 4 | 2 | 1 | 0 | 1 | 0 | 182/200 | 181/182 | 57/182 |
| 1 self-reported dose | 151 | 14 | 0 | 22 | 102 | 13 | 91.0% | 99.5% | 31.3% |
| 2 self-reported doses | 42 | 2 | 0 | 3 | 32 | 5 |  |  |  |
| 3 self-reported doses | 3 | 0 | 0 | 1 | 0 | 2 |  |  |  |
| Verified status (total) | 200 | 18 | 1 | 26 | 135 | 20 |  |  |  |
| **Women living with HIV, non-perinatally acquired** |  |  |  |  |  |  |  |  |  |
| Not vaccinated by self-report | 1 | 0 | 0 | 0 | 1 | 0 | 107/122 | 106/107 | 30/107 |
| 1 self-reported dose | 93 | 13 | 0 | 11 | 63 | 6 | 87.7% | 99.1% | 28.0% |
| 2 self-reported doses | 25 | 2 | 0 | 3 | 17 | 3 |  |  |  |
| 3 self-reported doses | 3 | 0 | 0 | 1 | 0 | 2 |  |  |  |
| Verified status (total) | 122 | 15 | 0 | 15 | 81 | 11 |  |  |  |
| **Women living with HIV, perinatally acquired** |  |  |  |  |  |  |  |  |  |
| Not vaccinated by self-report | 3 | 2 | 1 | 0 | 0 | 0 | 75/78 | 75/75 | 27/75 |
| 1 self-reported dose | 58 | 1 | 0 | 11 | 39 | 7 | 96.2% | 100% | 36.0% |
| 2 self-reported doses | 17 | 0 | 0 | 0 | 15 | 2 |  |  |  |
| 3 self-reported doses | 0 | 0 | 0 | 0 | 0 | 0 |  |  |  |
| Verified status (total) | 78 | 3 | 1 | 11 | 54 | 9 |  |  |  |

\*Of the 21 women without verification, 17 resided in a different country during primary school, 3 resided in remote areas of Botswana for which travel during this study was not possible, and 1 had records searched but information was not sufficient to verify vaccination.

Of the 429 participants with verified vaccination records, 248 (58%) had records located in the vicinity of Gaborone, and 181 (42%) had records located during travel outside of Gaborone. The majority, 62%, of vaccination records were health facility records, and 35% were school records; the remainder of vaccinations were verified using vaccination cards or medical records. Most vaccinated girls (n=315/429, 73%) initiated vaccination at school, and of these, most (n=303/315, 96%) received all doses at school; 12 received subsequent doses at a health facility. A minority (n=96/429, 22%) initiated vaccination at a health facility; of these, 93 (97%) received all doses at a health facility.

Agreement between self-report and verified ≥1 dose vaccination was >99% overall and in all groups (Table 1). One participant reported being unvaccinated, but records showed that she received 2 doses. Percent agreement by number of doses was 37% overall. By group, women without HIV (41%) and WLHIV-PA (36%) had the highest agreement regarding number of doses, and WLHIV-nPA (28%) had the lowest agreement. Underreporting number of doses was more common than overreporting. Of the 344 participants confirmed to have received 2 doses, 227 (66%) self-reported receiving 1 dose, and of 32 participants confirmed to have received 3 doses, 28 (88%) self-reported receiving 1 or 2 doses. Most participants who self-reported receiving 1 dose were confirmed to have received 2 doses (n=227/285, 80%).

Several challenges were encountered during the vaccination verification effort. Research assistants traveled >1000 km by car and flew to two locations. At health facilities and schools, storage and organization of records from several years prior was variable. Research assistants spent significant time searching on-site for student records and vaccination ledgers. Some road hazards were encountered reaching the diversity of locations nationwide, especially during the rainy season.

## DISCUSSION

We found that 99% of young adult women in Botswana who were mainly vaccinated as part of Botswana’s initial multi-age HPV vaccination cohort targeting 9–13-year-olds in 2015 accurately reported receiving ≥1 dose of HPV vaccine. However, the exact number of vaccine doses received was reported less accurately, demonstrating poorer recall of the details of their specific HPV vaccination history. Most often, participants underreported the number of doses received.

Our vaccination verification study demonstrated that it is possible, although resource-intensive, to find individual HPV vaccination records at schools and health facilities, including dates and number of doses, from a school-based vaccination programme. In our study of women aged 18–22 years, child/adolescent vaccination records were located for 95%. Nearly all (>99%) who self-reported receipt of ≥1 dose of HPV vaccine were verified to have been vaccinated. However, accuracy of exact number of self-reported doses compared to verification records was low (37% overall).

Research on vaccine impact and effectiveness requires accurate data on vaccination coverage, prior vaccination status and number of doses received (7, 8). In some countries, nationwide registry linkage enables research and programme monitoring with a high degree of confidence in immunization histories (9, 10). In the absence of national registries and record linkage, the challenges of obtaining accurate adolescent HPV vaccination histories have been noted in a variety of settings (11, 12, 13). A single-dose vaccine impact and effectiveness study in South Africa conducted among grade 10 vaccinees used self-reported vaccination status at age 17–18 years and followed up with medical record review and study vaccination registers, using probabilistic matching techniques; agreement among data sources was not reported (11). In the United States, which has no national vaccination registry, studies are often conducted within health systems in which individuals are continuously enrolled for several years (12). In another U.S. project, the five-site surveillance programme HPV-IMPACT uses medical record review and state Immunization Information System searches; vaccination remains unknown for half of precancer cases in a vaccine-eligible age group (13).

Many vaccination validation studies report test measures such as sensitivity, specificity, positive predictive value, negative predictive value, and Kappa statistic. Due to the small number of unvaccinated persons in this study, we determined that these measures would not be informative. Studies conducted in several high-income countries have found limitations to the validity of parent- and self-reported HPV vaccination, although most have not evaluated dose-specific agreement (14-17). One U.S. survey provided some data by number of doses: sensitivity and specificity of self/parent-reported receipt ≥1 HPV vaccine dose were ≥83% compared to medical records, but self-report of 3 doses was less accurate (14),.

Global data on vaccination coverage in Botswana indicated first-dose coverage of 76% in 2015 (15). Data from this study of participants recruited from UB and Gaborone area clinics suggests even higher ≥1 dose coverage from the initial multi-age cohort. While participants were unlikely to be representative of all young women in Botswana, they did include participants from all areas of the country and included women from different backgrounds. Coverage was likely impacted by a global HPV vaccine shortage during some years (16) that caused delays in vaccination in some cohorts. Our study suggests that coverage among girls who eventually enrolled at UB was extremely high in the initial multi-age cohort – with 100% self-reporting vaccination and nearly all having ≥1 dose verified. This study also suggests high coverage among WLHIV, a group at higher risk of cancers caused by HPV, including cervical cancer.

This study demonstrated high validity of self-reported ≥1-dose HPV vaccination among young women in Botswana, although recall of the exact number of doses received was less accurate. Most vaccinations were given in schools, but records were more easily found at nearby health facilities, reflecting the role of the health care system in school-based vaccination in Botswana. Although record-keeping methods varied, records do exist, and it was possible to verify vaccination history for most female study participants. However, verification was not possible for a subset of women, particularly for women who had immigrated as adolescents or young adults. Self-reported number of doses was unreliable; this is important information for consideration of self-reported vaccination history in studies of vaccine outcomes by number of doses.

Botswana has other national registries, including an HIV treatment registry. A national vaccination registry could capture accurate individual- and population-level vaccination data that could facilitate planning, delivery and evaluation of a vaccination programme to ensure fair vaccine distribution and coverage. While a national registry would not help with verifying coverage of young women who were vaccinated in another country before immigrating to Botswana, routinely collecting and reporting on HPV vaccination data nationally would help with determining coverage in the more remote areas of Botswana that were not reached in this study. Furthermore, an HPV vaccination registry would facilitate HPV-related research in Botswana that could provide evidence on vaccine effectiveness and support national immunization goals.

## Data Availability

Anonymised data from this study are available on request from the first author

## ACKNOWLEDGEMENTS

We acknowledge with thanks the contributions of Kereng Molly Rammipi, Kagiso Ndlovu, and Sarah Brewer.

## Data

Anonymised data from this study are available on request from the corresponding author.

## Authors’ Contributions

DRM: conceptualization, investigation, methodology, project administration, resources, supervision, visualization, writing original draft, reviewing and editing; AM: conceptualization, investigation, methodology, project administration, supervision, visualization, writing original draft, reviewing and editing; MM: investigation; MM: investigation; SP: investigation; JO: Writing – review & editing; RML: conceptualization, data curation, investigation, methodology, visualization, writing, reviewing and editing, software, supervision CM: writing-reviewing and editing; RL: writing-reviewing and editing; LEM: supervision; reviewing and editing, funding acquisition, conceptualization; JWG: conceptualization, formal analysis, methodology, project administration, resources, supervision, visualization, Writing - original draft, Writing – reviewing and editing, software, funding acquisition.

